# Development and clinical validation of a cross-sex translator of ECG drug responses

**DOI:** 10.1101/2024.12.27.24319698

**Authors:** Roshni Shetty, Stefano Morotti, Vladimír Sobota, Jason D. Bayer, Haibo Ni, Eleonora Grandi

## Abstract

Sex differences in cardiac electrophysiology are a crucial factor affecting arrhythmia risk and treatment responses. It is well-documented that females are at a higher risk of drug-induced Torsade de Pointes and sudden cardiac death, largely due to longer QTc intervals compared to males. However, the underrepresentation of females in both basic and clinical research introduces biases that hinder our understanding of sex-specific arrhythmia mechanisms, risk metrics, disease progression, treatment strategies, and outcomes. To address this problem, we developed a quantitative tool that predicts ECG features in females based on data from males (and vice versa) by combining detailed biophysical models of human ventricular excitation-contraction coupling and statistical regression models. We constructed male and female ventricular tissue models incorporating transmural heterogeneity and sex-specific parameterizations and derived pseudo-ECGs from these models. Multivariable lasso regression was employed to generate sets of regression coefficients (a cross-sex translator) that map male ECG features to female ECG features. The predictive ability of the translator was evaluated using an independent dataset that simulates the effects of various drugs and pharmacological agents at different concentrations on male and female models. Furthermore, we demonstrated a proof-of-concept clinical application using ECG data from age-matched subjects of both sexes under various drug regimens. We propose our cross-sex ECG translator as a novel digital health tool that can facilitate sex-specific cardiac safety assessments, ensuring that pharmacotherapy is safe and effective for both sexes, which is a major step forward in addressing disparities in cardiac treatment for females.

**One Sentence Summary:** We used biophysical and regression models for predicting ECG features across sexes to address disparities in sex-specific cardiac drug responses

## INTRODUCTION

Intrinsic biological differences in cardiovascular structure and function between males and females directly impact the likelihood, severity, and type of cardiovascular disease (*1, 2*). Underlying sex-related differences in ion-channel electrophysiology manifest at the scale of whole-heart electrocardiograms (ECG) (*3, 4*). Bazett first reported sex- and age-driven differences, noting that pre-menopausal females had prolonged QT intervals compared to age-matched male subjects (*5*), despite a smaller heart size than males. This is mainly attributed to a reduced gene expression for a wide variety of K^+^ channel subunits (including hERG) in female vs. male ventricles (*6, 7*). Female ECGs also exhibit shorter RR and QRS, and smaller T-wave amplitudes and ST angles than males (*8*).

The longer corrected QT duration due to reduced repolarization reserve makes females more susceptible to drug-induced Long-QT syndrome (LQTS) and torsades de pointes (TdP), which can trigger ventricular fibrillation (VF) and sudden cardiac death (*1, 4, 9–12*) and is a leading cause of drug attrition in the drug development pipeline. Safety guidelines mandate the use of two main metrics to assess the likelihood of drug-induced TdP for any new compound: in vitro studies to quantify hERG channel block and in vivo ECG studies to measure QT interval prolongation. While the validity of these metrics of TdP risk is the subject of much debate (*13*), an additional challenge is that females are underrepresented in clinical trials, leading to potential sex bias in these screens. This could render new compounds potentially ineffective or even harmful in vulnerable (e.g., diseased or aged) female populations (*11, 14–17*). Indeed, the approval of most drugs based on clinical trials predominantly conducted in males has likely contributed to overmedication of females (*18*). Thus, there is therefore an urgent need for tools that quantitatively evaluate sex-differences and improve drug-induced arrhythmia risk prediction for both sexes.

Computational modeling, grounded in experimental data, has proven powerful in dissecting key mechanisms underlying cardiac electrophysiology and arrhythmogenesis (*19, 20*), and invaluable in predicting drug cardiotoxicity (*21, 22*). By offering a framework that overcomes the limitations of species differences inherent in animal studies, computational modeling can provide an accurate representation of human cardiac physiology. Furthermore, simulations are cost-effective, scalable, and align with ethical standards by reducing reliance on animal testing. Incorporating sex differences into these models allows in-silico drug screening and prediction of electrophysiological features in a sex-specific manner (*17, 23–25*). Previous work from our team used data-driven models with biophysical simulations to translate (i.e., predict) action potential (AP) and Ca^2+^ transient (CaT) biomarkers in females given male data (and vice versa) (*26*). However, translation of ECG features is crucial for clinical application. To achieve this, we propose a multi-scale computational approach to translate ECG features across sexes, bridging cellular AP to the ECG.

We simulated male and female pseudo-ECGs using populations of male and female ventricular one-dimensional cable models. Multivariable lasso regression was employed to generate sets of regression coefficients (a cross-sex translator) that map male ECG to female ECG features. The predictive capability of the translator was assessed using an independent dataset that simulated the effects of various drugs and pharmacological agents at different concentrations on male and female models. Additionally, we demonstrated a proof-of-concept clinical application using ECG data from age-matched male and female subjects under various drug regimens (*27–29*). We propose that our cross-sex ECG translator could serve as a novel digital health tool for sex-specific cardiac safety assessments, promoting safe and effective pharmacotherapy for both sexes and helping to address existing treatment disparities.

## RESULTS

### Development and cross validation of the male-to-female ECG translator

We constructed male and female one-dimensional cable models (**Fig. 1A – Step 1**) of ventricular transmural tissue by simulating strands of myocytes with the O’Hara-Rudy cellular model (*30*), updated to integrate experimental data on sex differences in ion channels and Ca^2+^ handling (*6, 17, 31*) and their transmural variations (*30, 32*). Populations of 750 male and female cables were generated by random perturbation of baseline model parameters (*33*) and pseudo-ECGs were calculated (see Materials and Methods for details) (**Fig. 1A -Step 2**). For each model variant, we computed key ECG features: QRS duration (ORSdur), QT interval (QTint), T-wave amplitude (T-wave amp), T-peak-to-end duration (T-peak-end dur) when pacing at a basic cycle length of 1,000 ms (equivalent to a heart rate of 60 beats per minute). Notably, our models reproduce clinically observed sex differences, including longer QT interval and smaller T-wave amplitudes in females (*8*) (**Fig. 1B**). We then built the cross-sex translator by using lasso regression to yield a set of regression coefficients mapping male-to-female features (and vice versa) (**Fig. 1A – Step 3**). Given a male ECG, the corresponding female ECG features can be predicted as a function of the male ECG features (and vice versa) (**Fig. 1A -Step4**). We validated the ability of the translator to predict simulated female ECG features from given simulated male ECG features using an independent test set (219 male and female cables), not used to derive the regression model, and revealed an R^2^ > 0.94 for the four features when comparing actual vs. predicted values (**Fig. 1C**).

**Fig. 1.**
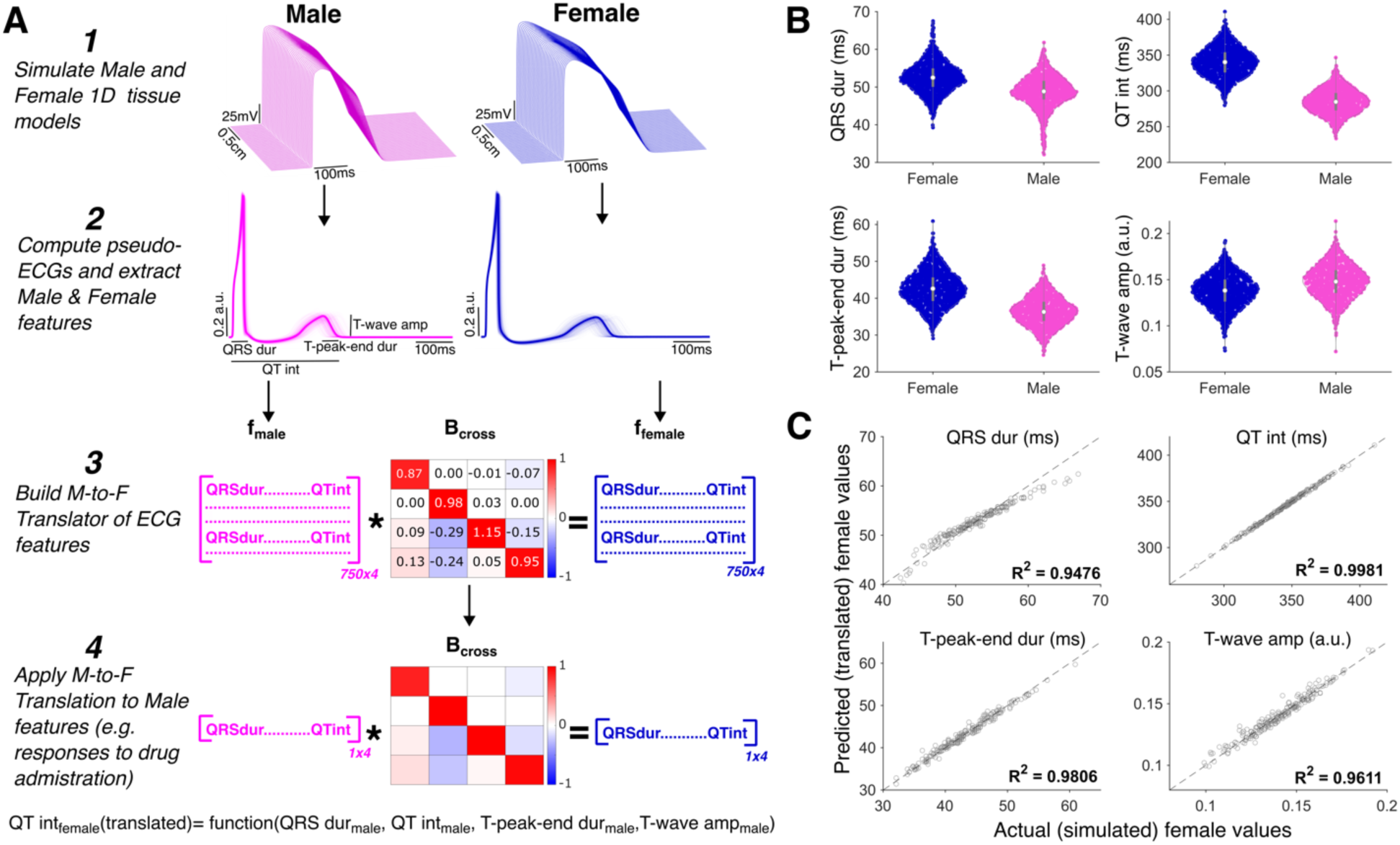
Development and validation of the cross-sex ECG translator. (A) Development workflow: *Step 1*. Male and female cable models of ventricular tissues are built to simulate propagating action potentials. ***Step 2*.** Pseudo-ECGs are computed for the male and female populations to extract ECG features (QRS duration (QRS dur), QT interval (QT int), T-wave amplitude (T-wave amp), T-peak-to-end duration (T-peak-end dur)). ***Step 3***. Lasso regression is used to predict the female ECG features based on the male ECG features, yielding B_cross_, a set of regression coefficients mapping male to female ECG features (vice versa is also possible). ***Step 4*.** B_cross_ is applied to predict ECG features in the output sex (“Translated female features”), given the values observed in the input sex (“Actual male features”), **(B)** Distribution of pseudo-ECG features from populations of male and female tissue models (n = 969 for each sex), **(C)** Cross-validation with an independent test set (n = 219 models for each sex). For each feature, the female values predicted by the translator are plotted against the actual (simulated) values.

### Male-to-female translator application and validation with simulated drug responses

To test the applicability of the ECG translator, we attempted to predict the concentration-dependent drug responses in female ECG features from the measured effects on the male pseudo-ECGs. We utilized a broad simulated dataset comprising 98 formulations of compounds (including anti-arrhythmic and other miscellaneous pharmacological agents)(*17, 34–36*). An example illustrating female ECG features that were predicted by the translator from simulated male ECG features in the presence of dofetilide, quinidine, ranolazine, and verapamil at their respective effective therapeutic plasma concentration (ETPC) are shown in **Fig. 2A**. The translator successfully predicted the simulated drug-induced effects in female ECG features when simulated male ECG features in response to the same drug were given as input, with an average <5% discrepancy between all four predicted and simulated female ECG features across all the 98 formulations in simulated datasets (**Fig. 2B**). Translation errors for individual features across 1-4x ETPC were as follows: QRS dur (0.63 ± 1.28 %); QT int (0.10 ± 0.15 %); T-wave amp (4.26 ± 9.84 %); T-peak-end dur (1.75 ± 2.71 %).

**Fig. 2.**
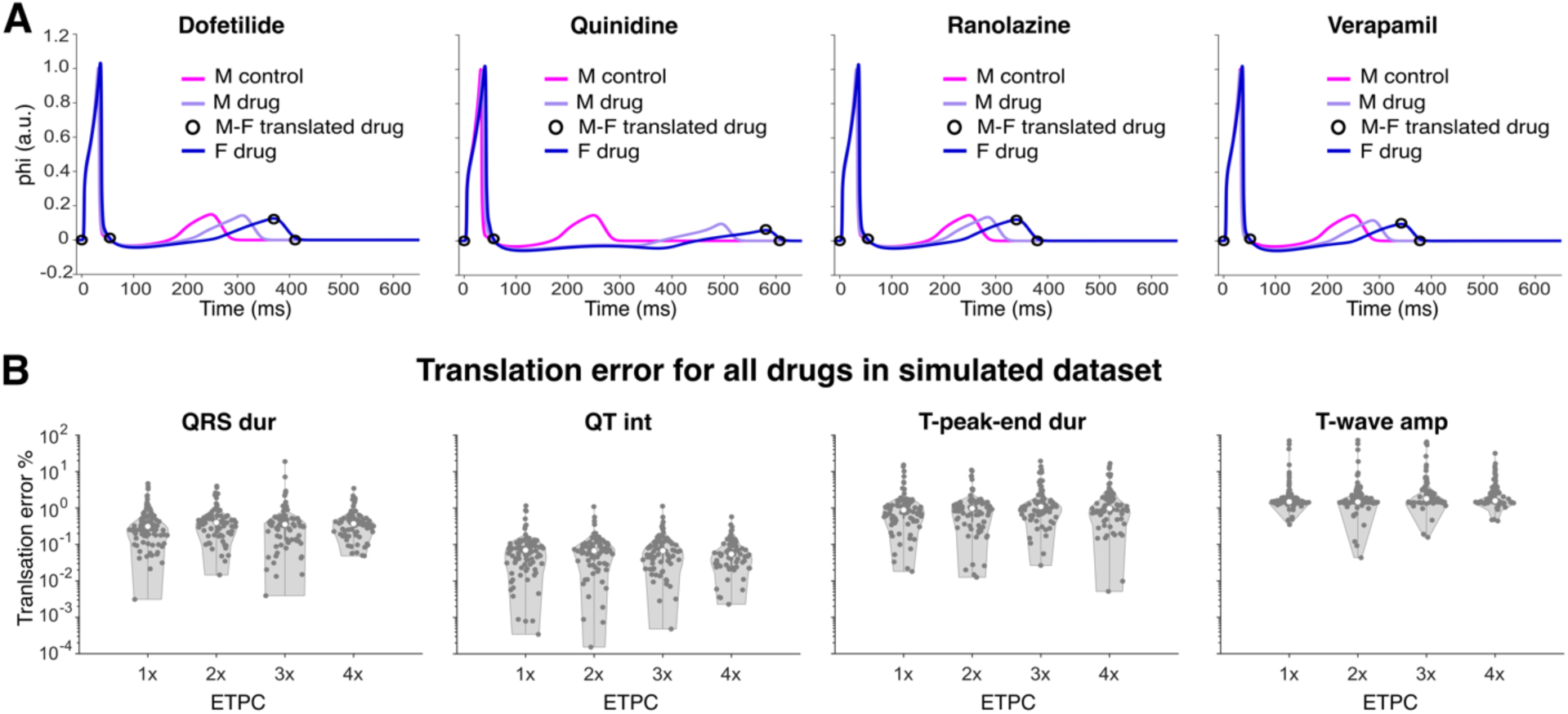
Translator application and validation with simulated drug responses. **(A)** The translator was validated against simulated pseudo-ECG drug responses. As an example, simulations of male and female pseudo-ECG under the action of dofetilide, quinidine, ranolazine, and verapamil at their respective effective therapeutic plasma concentration (ETPC) are shown. **(B)** Violin plots depicting the relative male-to-female translation error calculated as abs(1 - translated value/actual value) for translation of each feature under the action of 98 drug formulations at 1-4× ETPC. Data is excluded from analysis for cables without propagating action potentials and without positive T-waves under drug block (1×: 6, 2×: 16, 3×: 21, 4×: 27). Note the logarithmic scale on y-axis for % errors.

### Translator application and validation with clinical ECG data

To demonstrate proof-of-concept application of the translator to clinical data, we further applied the translator to 24-hour clinical ECGs from two different cohorts comprising healthy age-matched subjects of both sexes receiving the same dose of various anti-arrhythmic drugs (Dataset 1: Johannesen *et al.,* 2014 (*27, 29*)) and drug combinations (Dataset 2: Johannesen *et al.,* 2016 (*28, 29*)) (**Fig. 3 and 4**). The female features predicted by translating (shaded black in **Fig. 3A** and **Fig. 4A**) the male data (shaded pink in **Fig. 3A** and **Fig. 4A**) matched closely the actual female data (shaded blue in **Fig. 3A** and **Fig. 4A**). The translator predicted drug-induced relative changes from the baseline in female ECG features from the corresponding male ECG data with an average translation error <6% across all four features for all timepoints in both Dataset 1 (**Fig. 3B**) and Dataset 2 (**Fig. 4B**). The translation errors for individual features comprising Datasets 1 and 2 were as follows: QRS dur (1.42 ± 1.19 %); QT int (0.97 ± 0.82 %); T-wave amp (5.24 ± 4.78 %); T-peak-end dur (5.26 ± 4.82 %). Dofetilide was administered with different initial concentrations in the two studies, thus leading to different levels of QTc prolongation. Notably, the translator demonstrates reasonable performance across both datasets, achieving an average error for dofetilide of 6.92 ± 7.08% in Dataset 1 and 2.68 ± 2.58% in Dataset 2 across all four features.

**Fig. 3.**
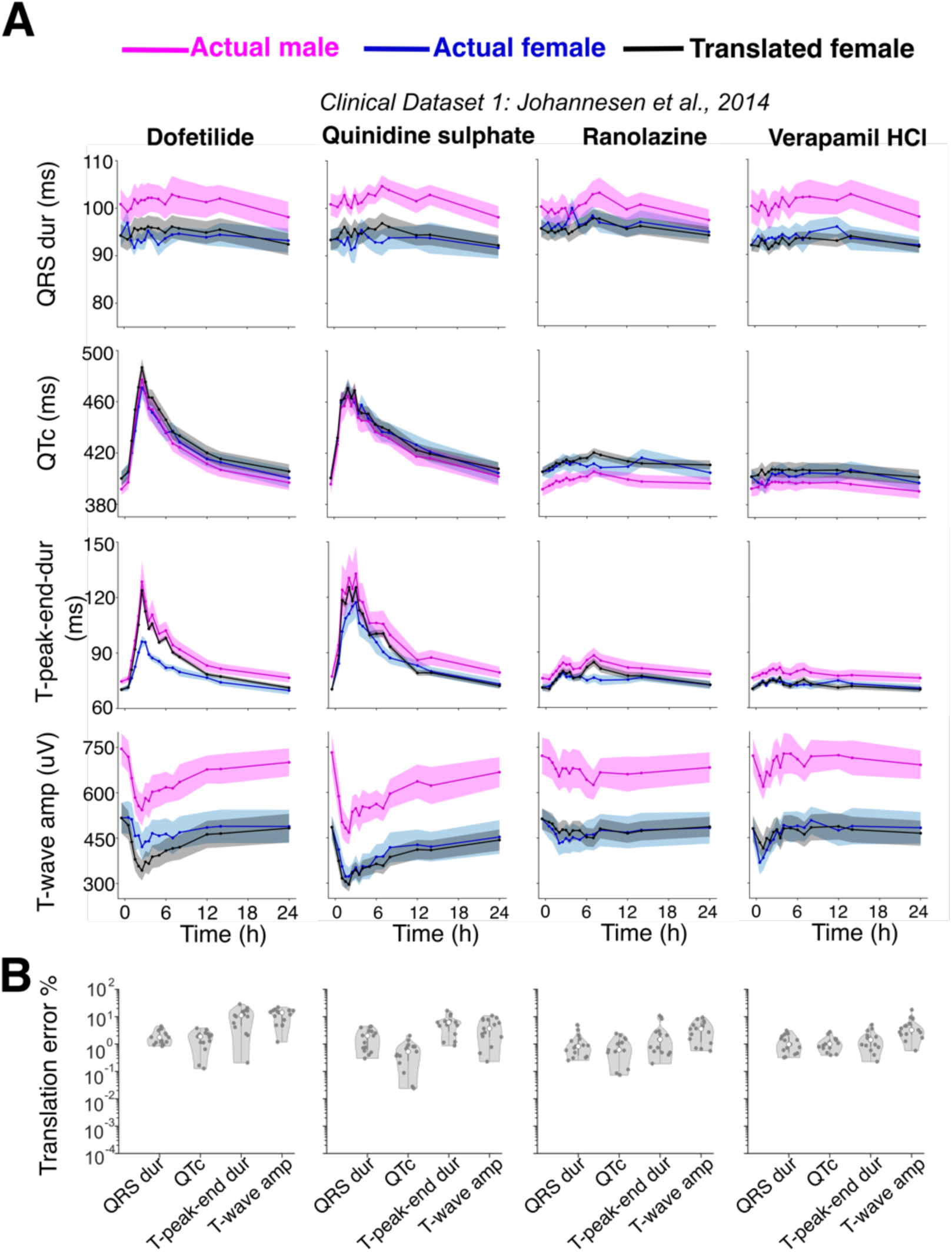
Translator application and validation with clinical ECG data comprising single drug responses. **(A)** The translator performance was tested for prediction of female ECG features based on male clinical ECGs using clinical dataset 1 (*27*); Row 1. QRS duration; Row 2. QTc interval; Row 3. T-peak-end duration; Row 4. T-wave amplitude. Male features (pink) from clinical ECG data under drug administration over a 24-hour period (solid lines indicate the mean value, and shaded area indicates the SEM) are translated into female responses (black). The latter are compared with clinical female features under the same drugs (blue). **(B)** Translation error for the drugs in **(A)**, calculated for the four features as abs(1-mean(translated value)/mean(actual value)) at each time point. Each column corresponds to a single drug administration as listed. Dataset 1(*27*) comprised participants aged 26.9 ±5.5 years (11 males, 11 females). All feature values were placebo corrected. Note the logarithmic scale on y-axis for % errors.

**Fig. 4.**
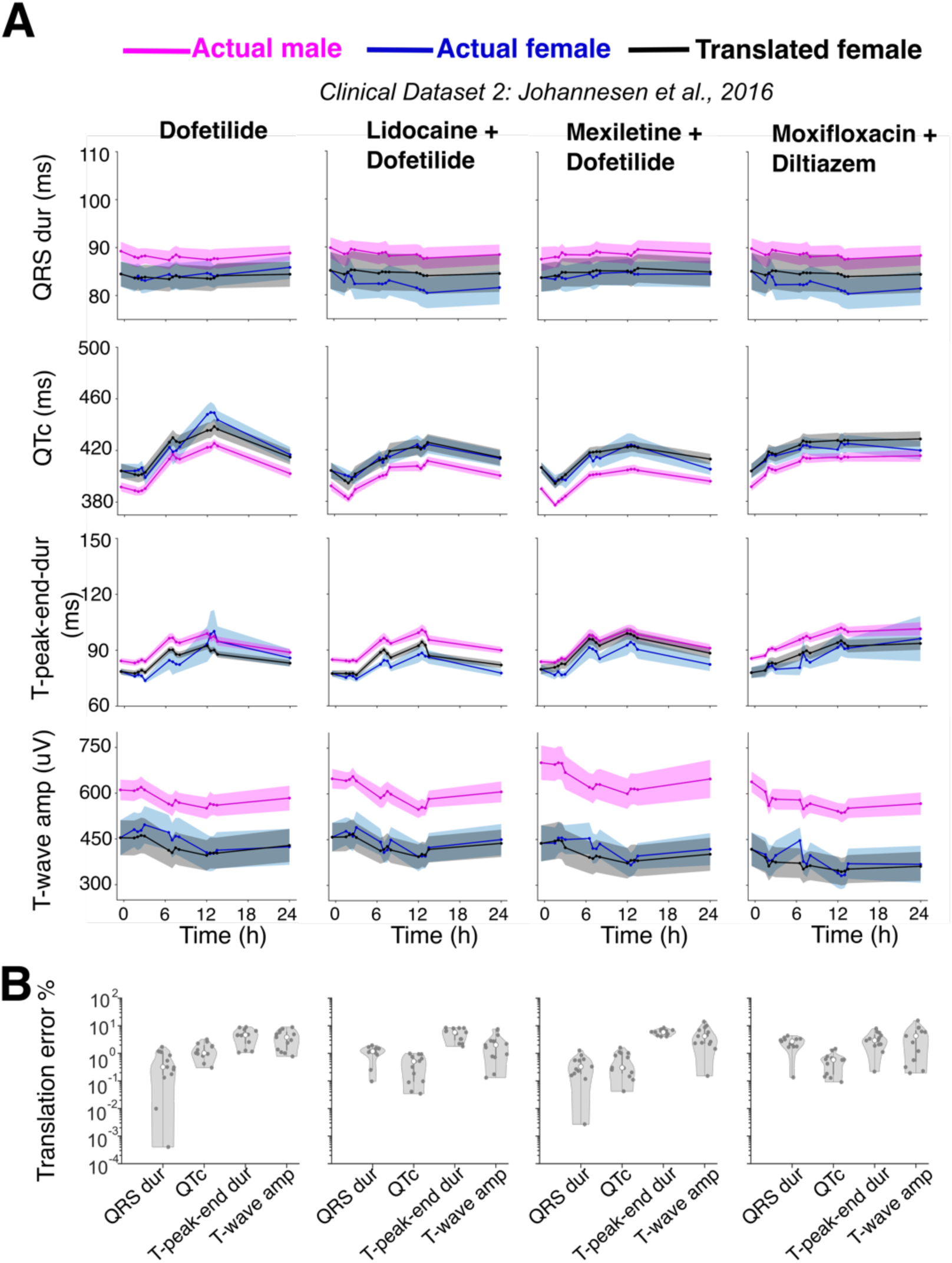
Translator application and validation with clinical data comprising drug combination responses. **(A)** The translator performance was tested for prediction of female ECG features based on male clinical ECGs using clinical dataset 2 (*28*); Row 1. QRS duration; Row 2. QTc interval; Row 3. T-peak-end duration; Row 4. T-wave amplitude. Male features (pink) from clinical ECG data under drug administration over a 24-hour period (solid lines indicate the mean value, and shaded area indicate the SEM) are translated into female responses (black). The latter are compared with clinical female features under the same drugs (blue), **(B)** Translation error for the drugs in **(A)**, calculated for the four features as abs(1-mean(translated value)/mean(actual value)) at each time point. Each column corresponds to a single drug administration as listed. Dataset 2 (*28*) comprised participants aged 26.1 ± 4.9 years (13 males, 9 females). All feature values were placebo corrected. Note the logarithmic scale on y-axis for % errors.

## DISCUSSION

### Summary and key outcomes

The underrepresentation of females in research remains a persistent challenge (*15*). This has led to gaps in our understanding of sex-specific biological differences and their impact on disease progression, treatment efficacy, and drug safety. As a result, females are often at higher risk of adverse drug reactions (*37, 38*) and suboptimal treatment outcomes due at least in part to the reliance on data predominantly derived from male subjects. To address this issue, we developed a quantitative tool combining biophysical modeling of human ventricular excitation-contraction coupling with statistical regression models to predict ECG features in females based on male data (and vice versa). Our team has previously demonstrated the success of multivariable linear regression in developing cross-species (*39*) and cross-sex translators of cellular action potential (AP) and calcium transients (CaT) biomarkers (*26*), which showed strong agreement between predicted and experimentally observed features. Building on this foundation, we employed lasso regression (*40*) for our cross-sex ECG translator. This method improves upon traditional regression techniques by eliminating redundant features and reducing feature co-dependency, thus yielding more accurate and interpretable models. We validated the tool using an independent dataset simulating the effects of various drugs and pharmacological agents at various concentrations on male and female models. Additionally, we showcased a proof-of-concept clinical application using ECG data from age-matched individuals of both sexes under different drug regimens. This study represents a significant technological innovation, introducing a much-needed health tool that predicts ECG features across sexes.

### Biological insights and applications in drug development and clinical settings

Our findings highlight the potential of our novel cross-sex ECG translator, validated using clinical data from male and female subjects, to serve as a powerful tool for bridging the knowledge gap in quantitatively predicting sex-specific ECG responses to drugs. The translator offers a practical application in drug discovery and clinical settings, enabling precise predictions of ECG features across sexes. For example, the translator could identify cases where QT interval prolongation remains within normal limits for males but may pose an arrhythmogenic risk in females, enabling these scenarios to be addressed with caution. A key strength of our approach is the integration of biophysical modeling with statistical learning techniques. This approach not only provides a mechanistic understanding of the physiological processes driving sex-specific ECG differences but also enables the discovery of emergent properties that can be extrapolated to new, previously untested conditions. For instance, biophysical simulations can be evaluated across different drug concentrations and traced back to identify parameters that influence outputs like biomarkers and arrhythmia susceptibility (*33, 41*). These capabilities are often limited when AI-based techniques are applied solely to clinical data, which may lack the underlying biological context and mechanistic insights that are essential for accurate predictions. As highlighted by previous studies, sex-specific simulations can uncover and predict differences in dose responses and arrhythmia susceptibility to pharmacological agents (*17, 23–25*). Conceptually, the translator also offers the ability to extract novel insights from existing clinical data. For example, by analyzing ECG feature changes at varying drug concentrations between sexes, it can provide valuable insights into dose-dependent effects and help identify sex-specific differences in drug metabolism or ion channel sensitivity.

The translator could also enable identification of new sex-specific metrics for arrhythmia prediction, leveraging a range of ECG features, not just QT intervals, as done in a recent study aimed at predicting sex-specific QTc prolongation and drug-induced proarrhythmic risk using cardiac emulators (*42*). This would provide a more comprehensive tool for assessing cardiac risk in both males and females, contributing to more personalized and precise healthcare. The cross-sex ECG translator could enhance the accuracy of cardiac safety evaluations for females, addressing the historical underrepresentation of females in clinical research and helping to mitigate sex-based disparities in medicine. We also envision the use of the translator as a valuable teaching tool, which could train clinicians to recognize the impact of drugs on ECGs in a sex-specific manner and enhance their ability to assess cardiac responses in males and females. Importantly, our work underscores the critical need for systematic consideration of sex as a biological variable in cardiovascular research.

### Limitations and Future Directions

#### Technical challenges and enhancements

During our drug simulations, we encountered challenges such as propagation failure due to limitations in the O’Hara-Rudy (*30*) sodium current (*I*_Na_) formulation. In some cases, the *I*_Na_ formulation may overestimate the drug effects and significantly disrupt excitation propagation, leading to inverted T-waves, which might not necessarily reflect clinical outcomes. To address these limitations, our future work will focus on incorporating alternative *I*_Na_ formulations and validation of the translator using other ventricular models, including the Grandi-Bers (*43*) and ToR-ORd (*32*) models. Additionally, our drug simulations currently rely on a simplified pore-block model that doesn’t account for state- or use-dependent blocks. If sex-dependent drug metabolism influences drug responses, we will integrate this complexity into our modeling framework to enhance the performance of our translator.

The current translator uses a 1D strand model for pseudo-ECG generation, offering computational efficiency and reasonable accuracy when compared to clinical data. However, this approach lacks the anatomical detail of 3D whole-heart models. Future work will evaluate the translator’s performance against 3D ventricular models (*44*) that incorporate apicobasal and transmural heterogeneity to improve predictive precision, particularly for complex conditions or drug responses. Another avenue for future work involves the systematic comparison of our translator’s performance against other AI-based tools designed for ECG analysis, such as deep learning models.

#### Heterogeneous patient cohorts

We evaluated the cross-sex ECG translator using a cohort of healthy young adults. Our findings highlight that male and female ECG features exhibit strong correlations under normal physiological conditions, as captured by the cross-sex regression model. This suggests shared patterns of relative changes in ECG features across sexes, which are maintained in young adults under normal conditions. However, these relationships may shift in aging and diseased states, where additional features might become significant, and may also require the adoption of non-linear or polynomial regression models. The translator’s applicability could be expanded to account for age, disease, and fluctuations in sex hormones by incorporating ECG data from a broader, more diverse set of clinical databases. This will involve simulating age- and disease-specific phenotypes within the biophysical model to create more robust, age- and disease-sensitive translators.

#### Sex-differences in pharmacokinetics

Our current translator development does not take into account potential sex-differences in pharmacokinetics, which can significantly influence drug absorption, distribution, metabolism, and elimination, ultimately affecting drug efficacy and safety across sexes (*45, 46*). This is a critical consideration, as identical drug dosages can lead to varying effective concentrations at the target site, resulting in differences in therapeutic outcomes and adverse effects. Addressing this issue is essential for improving the accuracy of clinical cardiac safety assessments, which should prioritize the influence of sex-based pharmacokinetics and pharmacodynamics. Our analysis revealed sex differences in plasma drug concentrations for quinidine and verapamil in the clinical studies we used (**Fig. S1-S2**), despite all subjects receiving identical doses. However, the drugs tested are potent enough that their ionic current block was only modestly affected by these variations. In the case of dofetilide from clinical dataset 1, while the relative QT prolongation is similar in male and female when simulating the same drug concentration, simulating the maximum drug concentration shows a 60% increase in male QTc vs. a 71% increase in female (**Table S1-S2**). In most cases, the translation model used here, which assumes similar perturbations of ion channels in males and females, remains valid. Nevertheless, accounting for pharmacokinetic differences could enhance the utility of the model in clinical applications, ultimately leading to better-informed, sex-specific dosing guidelines and safer therapeutic strategies.

## MATERIALS AND METHODS

### Male and female human ventricular models integrating experimentally identified sex and transmural differences

Previously implemented sex differences in protein expression of ion channels (*6*) in the O’Hara-Rudy model (*30*), served as the basis for our simulations. Compared to males, females exhibited reduced expression of K^+^ channel subunits such as HERG, minK, Kv1.4, KChIP2, as well as reduced expression of connexin 43 (*6*). In addition to K^+^ channel expression, work by our group has shown several Ca^2+^ handling proteins that regulate intracellular diastolic Ca^2+^ and CaT decay, may play a role in the predisposition of females to TdP (*17*). Here, we applied and updated these established sex-specific parametrizations of human ventricular myocytes incorporating transmural heterogeneity (*6, 17, 30, 31*). All adjustments (summarized in **Table 1**) were applied as scaling factors to ionic currents, fluxes, and transporters, except for I_Ks_. For I_Ks_, additional modifications included changes in the voltage dependence of steady-state activation (x_s1ss_) and (x_s2ss_) and their time constants (τ_xs1_ and τ_xs2_), as detailed in Equations 1-4 (*31*). **Fig. 5** shows the simulated male and female baseline models for the endocardial and epicardial cell types, incorporating the parameterization listed in **Table 1**.

**Table 1.** Sex-based differences in the human ventricle. Scaling factors are relative to the male endocardial <u>model. M – male; F – female; endo – endocardial cell; epi – epicardial cell.</u>

| Parameter | Definition | F vs M | Epi vs Endo | F endo scaling factor | M epi scaling factor | F epi scaling factor |
| --- | --- | --- | --- | --- | --- | --- |
| $G_{tos}$ | Maximal conductance of the slow component of the transient outward $K^+$ current | ↓ | ↓ | 0.64 | 0.6 | 0.26 |
| $G_{Kr}$ | Maximal conductance of the rapid delayed rectifier $K^+$ current | ↓ | ↑ | 0.80 | 1.86 | 1.49 |
| $G_{Ks}$ | Maximal conductance of the slow delayed rectifier $K^+$ current | ↓ | ↑ | 0.83 | 1.04 | 0.87 |
| $G_{K1}$ | Maximal conductance of the inward rectifier $K^+$ current | ↓ | ↓ | 0.86 | 0.98 | 0.74 |
| $G_{NaCa}$ | Maximal conductance of the $Na^+/Ca^{2+}$ exchange current | ↑ | ↑ | 1.15 | 1.1 | 1.27 |
| $G_{pCa}$ | Maximal conductance of the sarcolemmal $Ca^{2+}$ pump current | ↑ | = (F)<br>↓ (M) | 1.6 | 0.88 | 1.6 |
| $CMDN_{max}$ | Maximal buffering capacity of calmodulin | ↑ | ↑ | 1.21 | 1.07 | 1.41 |
| $Cx43 (R_i)$ | Intracellular resistance in the 1D cable (parameter values adjusted from Cx43 expression) | ↓ | ↓ | 0.68 | 0.94 | 0.61 |
| $G_{NaL}$ | Maximal conductance of the late $Na^+$ current | = | ↓ | 1 | 0.6 | 0.6 |
| $G_{to}$ | Maximal conductance of the transient outward $K^+$ current | = | ↑ | 1 | 2 | 2 |
| $P_{Ca}, P_{CaNa}, P_{CaK}$ | Permeability to $Ca^{2+}$, $Na^+$, $K^+$ through the L-type $Ca^{2+}$ channel | = | ↑ | = | 1.2 | 1.2 |
| $P_{NaK}$ | Permeability of the $Na^+/K^+$ ATPase current | = | ↓ | 1 | 1 | 0.94 |
| $G_{Kb}$ | Maximal conductance of the $K^+$ background current | = | ↓ | 1 | 0.6 | 0.6 |
| $J_{upNP}$ | Non-phosphorylated $Ca^{2+}$ uptake flux, via SERCA pump | = | ↑ | 1 | 1.42 | 1.42 |
| $J_{upCAMK}$ | CAMK phosphorylated $Ca^{2+}$ uptake flux, via SERCA pump | = | ↑ | 1 | 1.42 | 1.42 |

**Fig. 5.**
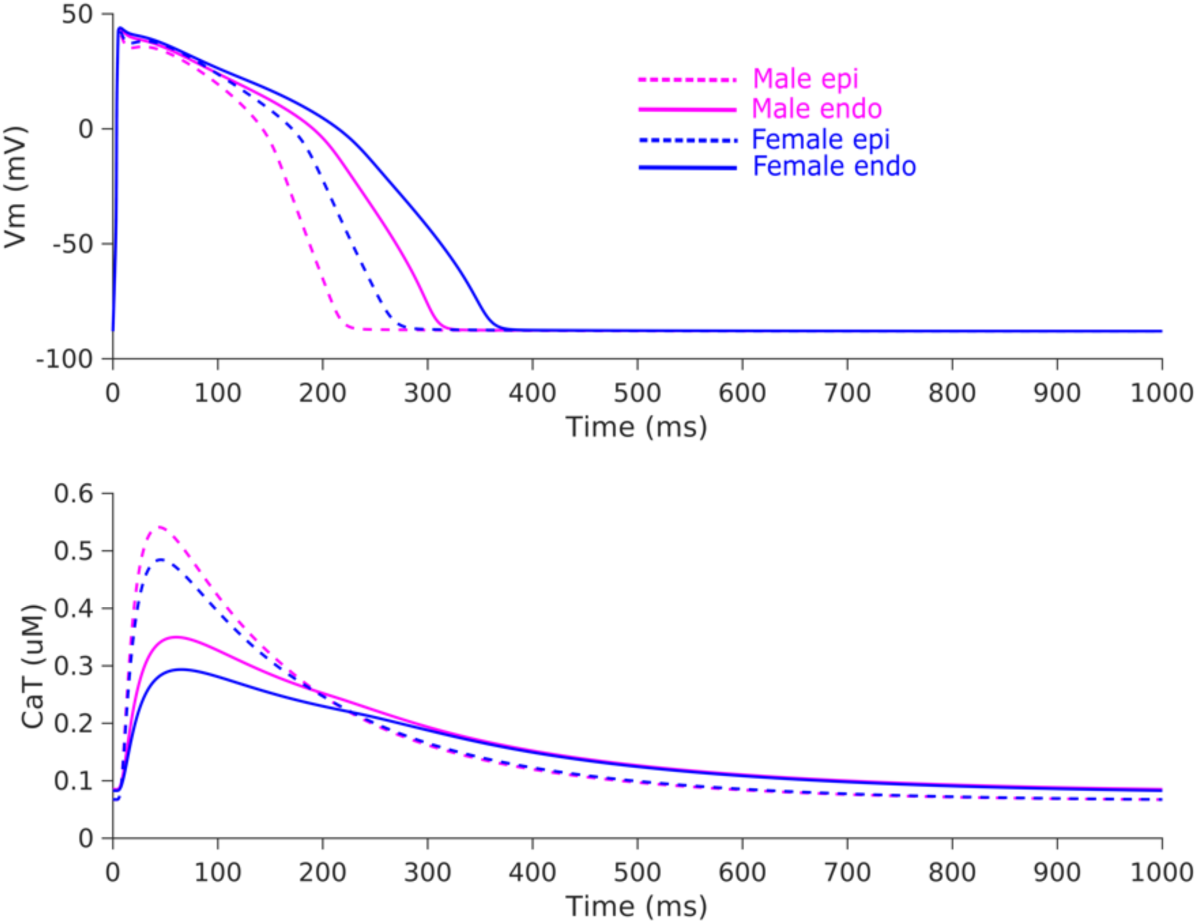
Simulated sex and transmural differences listed in **Table 1** in the O’Hara-Rudy single cell model. Top: Transmembrane voltage (*V*_m_); Bottom: Calcium transient (CaT)

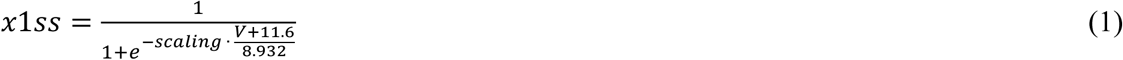

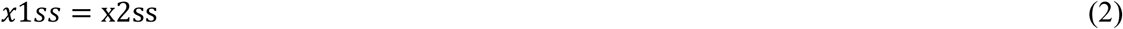

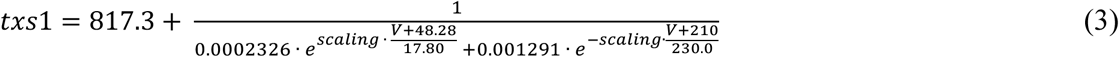

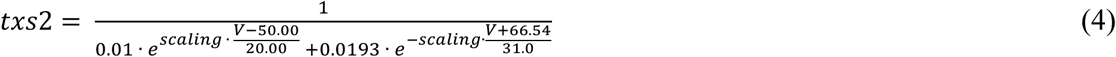

where scaling = 1 (M endo); 0.83 (F endo); 1.04 (M epi); 0.87 (F epi)

### One-dimensional cable models and pseudo-ECG calculation

The male cable comprised 205 cells and the female cable comprised 190 connected end to end by intracellular resistance. Female cable length was scaled to 90% of the male length to reflect smaller ventricular wall thickness in females vs males (*23*). In both male and female cable models, 50% of the cable contained endocardial cells and the other 50% epicardial cells, with a linear gradient applied to *G_Kr_* to simulate a gradual decrease in action potential duration (APD) from the endocardium to the epicardium, as previously simulated (*31*). The first 5 cells of the cable were stimulated from the endocardial end. The first 20 and last 20 cells were excluded for pseudo-ECG computation to avoid edge effects.

The pseudo-ECGs were computed by calculating the extracellular unipolar potentials Φ_e_ for the male and female population of cables according to Equation 5 (*47, 48*).

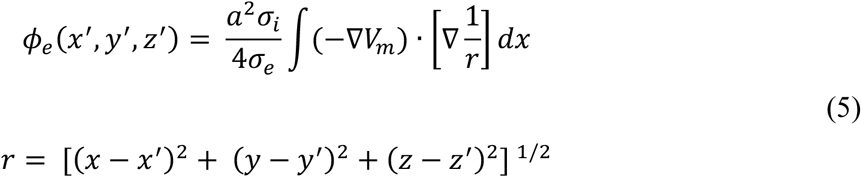

Where *V_m_* is the transmembrane potential, ∇*V_m_* is the spatial gradient of *V_m_*, *a* is the radius of the fiber, σ_i_ is the intracellular conductivity, σ_e_ is the extracellular conductivity, and *r* is the distance from a source point (x, y, z) to a field point (x’, y’, z’). Φ_e_ was computed at a virtual electrode located 2.0 cm away from the epicardial end of the cable. All pseudo-ECGs were normalized to the QRS amplitude of the baseline male pseudo-ECG.

### ECG feature calculation

For each pseudo-ECG, we calculated key ECG features, including QRS duration, QT interval, T-peak-to-end duration, and T-wave amplitude, using a custom software (**Fig 6**.). The ‘Q’ point was designated as the time at which the cable was stimulated from the endocardial end. The ‘R’ point was identified as the time when the maximum value of extracellular potential (Φ_e_) occurred. The ‘S’ point was determined as the time when Φ_e_ first crossed a value of 0.01 following the R peak. T-wave amplitude was measured as the maximum value of Φ_e_ following the S point. The end of the T wave was identified by finding the intersection of the line with the maximum rate of decay following the T-wave peak and the horizontal line at Φ_e_ = 0. QRS duration was calculated as the time interval between the Q and S points. The QT interval was calculated as the time interval from the Q point to the end of the T wave. T-peak-to-end duration was measured as the interval between the time of T-wave amplitude and the end of the T wave.

**Fig. 6.**
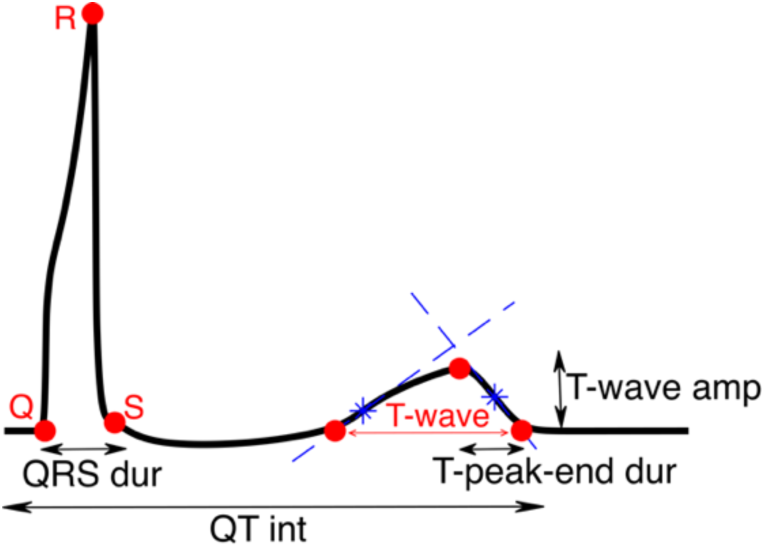
Calculation of pseudo-ECG features

### Population modeling and translator development

An initial population of 1,000 male and 1,000 female cable models was generated by random perturbation of model parameters (**Table 2**) using a log-normal distribution with a standard deviation of 0.1. From this population, 750 male and 750 female cables were used as the training dataset to develop the cross-sex translator “B_cross_” (see **Fig 1**. for schematic) employing lasso regression (*40*). The inbuilt MATLAB function ‘lasso’ was used with 10-fold cross-validation. Using 750 models for training yielded the same regression coefficients as using a larger number of models, so we chose not to increase the initial population size. From the initial populations, 219 male and female cables (not used to develop B_cross_) were used for validation and 31 models were excluded from analysis due to propagation failure and/or inverted T-waves.

**Table 2.**
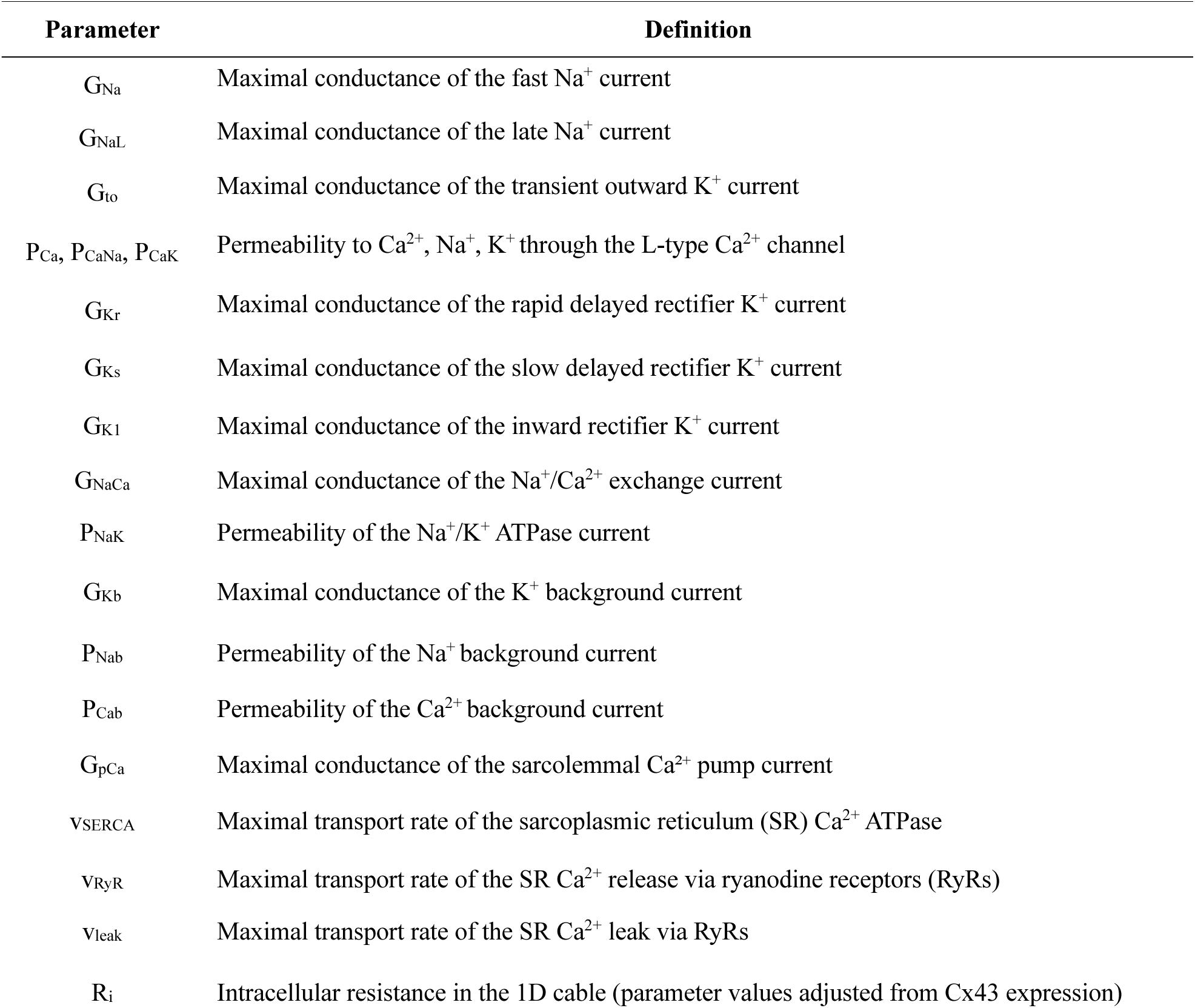
Model parameters randomly perturbed to introduce variability during population generation

### Drug simulations

Drug effects were simulated with male and female models to validate the applicability of the ECG translator to predict the concentration-dependent drug responses in female ECG features given the measured effects on the male pseudo-ECGs. We utilized a broad simulated dataset comprising 98 compounds (including anti-arrhythmic and other miscellaneous pharmacological agents) (*17, 34–36*). Each drug was simulated using a pore block model based on the available half-maximal inhibitory concentration (IC_50_) values and Hill coefficients for various ion channels. We simulated various drug concentrations, ranging from 1 to 4 times their effective therapeutic plasma concentration (ETPC), at 1 Hz pacing.

### Clinical ECG data processing

Dataset 1 and Dataset 2 were obtained from PhysioNet (*29*), where any feature value for each subject was reported in triplicate. We calculated the average of these triplicate values for each male and female subjects. QT values were rate-corrected using the formula:

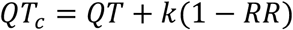

where k is the slope of the best-fit line for the relationship between QT and RR and QTc=QT at RR = 1 s. Note that QRS duration, T-peak-end duration, and T-wave amplitude were not rate-corrected. Placebo-corrected changes from baseline were applied to all feature values. For instance, the relative change in QTc at time ‘t’ from baseline ‘t_0_’was calculated as:

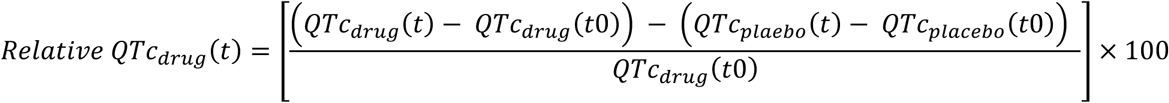

### Code availability

All codes used to perform simulations and data analysis were generated in MATLAB (MathWorks, Natick, MA, USA), version R2023b and Python3. Population-level simulations were performed with a computing cluster with Intel Xeon CPU E5-2690 v4 at 2.60 GHz 28 CPUs (56 threads) + 132 GB, and a standard laptop was used for data analysis. All source codes (and related documentation) and all simulated data used in this study are available for download at https://github.com/drgrandilab/

## Data Availability

All codes are available online at https://doi.org/10.13026/C2HP45

https://doi.org/10.13026/C2D016

## Funding

Institute of International Education Quad Fellowship (RS)

American Heart Association Career Development Award 24CDA1258695 (HN) American Heart Association Postdoctoral Fellowship 20POST35120462 (HN) NHLBI Grants R01HL131517, R01HL141214, and P01HL141084 (EG) NHLBI Grant R01HL170521 (EG, HN)

NHLBI Grants R00HL138160, R01HL171057, and R01HL171586 (SM)

National Institute of Aging Grant R03AG086695 (EG)

National Institutes of Health Stimulating Peripheral Activity to Relieve Conditions Grant 1OT2OD026580-01 (EG)

University of California Davis School of Medicine Dean’s Fellow Award (EG) France-Berkeley Fund (EG, JDB)

## Author contributions

Each author’s contribution(s) to the paper should be listed [we encourage you to follow the CRediT model]. Each CRediT role should have its own line, and there should not be any punctuation in the initials.

Examples:

Conceptualization: EG, HN, SM, RS

Methodology: EG, HN, SM, RS

Investigation: RS

Visualization: RS

Funding acquisition: RS, EG

Project administration: EG

Supervision: EG, HN

Writing – original draft: RS

Writing – review & editing: EG, HN, SM, JDB, VS

## Competing interests

Authors declare that they have no competing interests.

## Data and materials availability

All data needed to evaluate the conclusions in the paper are present in the paper and/or the Supplementary Materials. Data and source codes are freely available for download at: https://github.com/drgrandilab/

## Notes

### Competing Interest Statement

The authors have declared no competing interest.

### Author Declarations

The study used only openly available human data that were originally located at https://doi.org/10.13026/C2HP45 and https://doi.org/10.13026/C2D016

