## Supplementary material for "Development and clinical validation of a cross-sex translator of ECG drug responses": Data Supplement

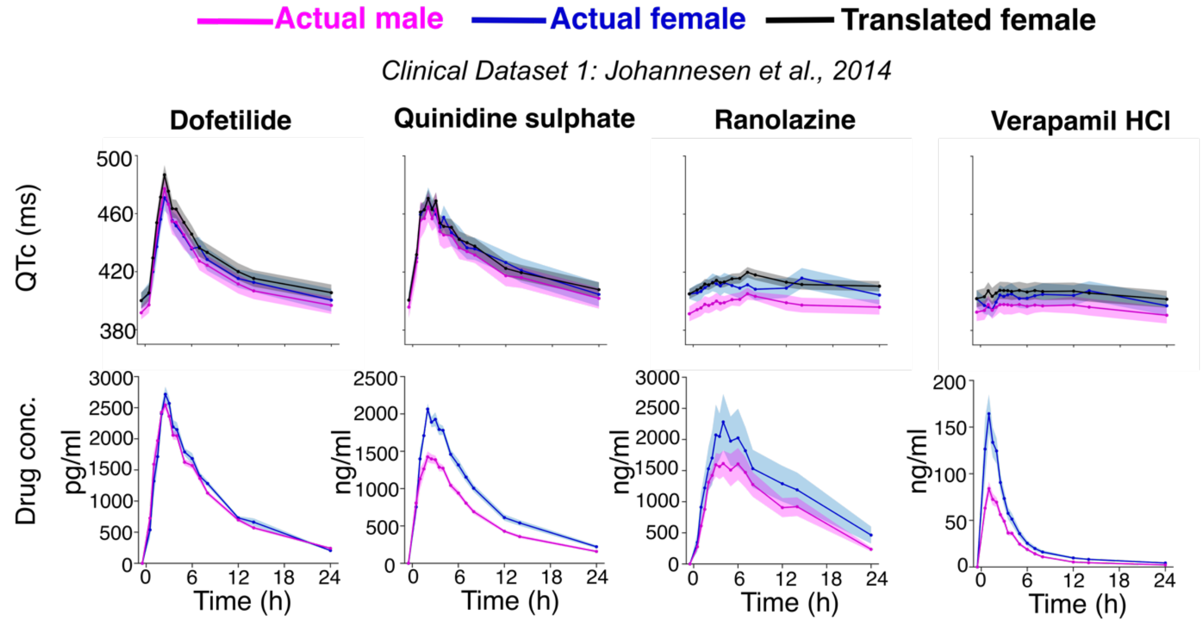


**Fig. S1.** Drug plasma concentrations (bottom) vs time and corresponding QTc intervals (top; re-plotted from Fig. 3A) over time for males (pink) and females (blue) in clinical dataset 1 *(27, 29*). Solid lines represent the mean values, with shaded areas indicating the standard error of the mean (SEM), and each column corresponds to a separate drug administration and monitoring over a 24-hour period. All subjects received identical drug dosages prior to ECG monitoring during a particular drug evaluation *(27)*.

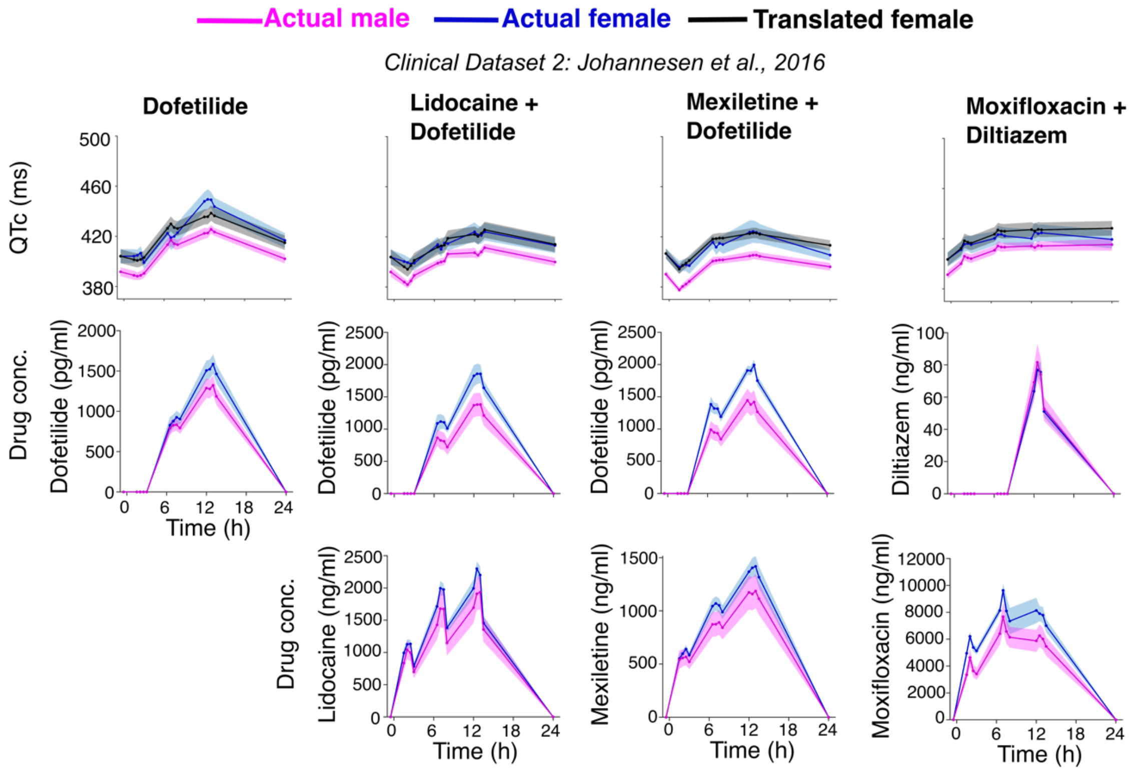


**Fig. S2.** Drug plasma concentrations (bottom) vs time and corresponding QTc intervals (top; re-plotted from Fig. 4A) over time for males (pink) and females (blue) in clinical dataset 2 *(29, 28)*; Solid lines represent the mean values, with shaded areas indicating the standard error of the mean (SEM), and each column corresponds to a separate drug administration and monitoring over a 24-hour period. All subjects received identical drug dosages prior to ECG monitoring during a particular drug combination evaluation and were dosed three times during the day *(28)*.

**Table. S1. Effects of sex-diverging** plasma concentrations of quinidine on major ionic currents at maximum value of difference in mean plasma concentrations during quinidine administration in male vs female: 2065 ng/mL (2638 nM) in female vs 1425 ng/mL (1820 nM) in male at time point 2 hours (shown in **Fig. S1**). Row 1: IC50 (nM) and hill coefficient ($n_h$) for ion channels blocked by quinidine (Data from *(34)*); Effect k = $1/(1+([D]/IC50)^{nh})$, where [D] is the drug concentration at 1820 nM (Row 2) and 263 8nM (Row 3).

| Quinidine CIPA | $I_{Kr}$ | $I_{NaL}$ | $I_{CaL}$ | $I_{Na}$ | $I_{to}$ | $I_{K1}$ | $I_{Ks}$ |
|---|---|---|---|---|---|---|---|
| IC50 ($n_h$) | 992 (0.8) | 9417 (1.3) | 51592.3 (0.6) | 12329 (1.5) | 3487.4 (1.3) | 39589919 (0.4) | 4898.9 (1.4) |
| k [D] = 1820 nM | 0.3810 | 0.8944 | 0.8815 | 0.9463 | 0.6996 | 1 | 0.80 |
| k [D] = 2638 nM | 0.3138 | 0.8395 | 0.8562 | 0.9100 | 0.5898 | 0.9755 | 0.7041 |

**Table. S2.** Analysis of simulated QTc from baseline male and female cable models and following administration of quinidine concentration 1820 nM (Row 1) and 2638 nM (Row 2).

| Quinidine Concentration [D] | QTc simulated (male) ms | QTc simulated % increase from baseline male (285.3ms) | QTc simulated (female) ms | QTc simulated % increase from baseline female (340.8ms) |
|---|---|---|---|---|
| 1820 nM | 456.0 | 59.8% | 533.4 | 56.5% |
| 2638 nM | 498.6 | 74.8% | 582.0 | 70.8% |
